# The use of the positive deviance approach for healthcare system service improvement: A scoping review protocol

**DOI:** 10.1101/2023.11.05.23298127

**Authors:** Ayelign Mengesha Kassie, Elizabeth Eakin, Biruk Beletew Abate, Aklilu Endalamaw, Anteneh Zewdie, Eskinder Wolka, Yibeltal Assefa

## Abstract

**Introduction:** Healthcare systems are currently facing challenges in enhancing access and improving the quality of healthcare services around the world, and one of the innovative strategies that have been utilized to address such challenges is the positive deviance (PD) approach. The approach assumes that identifying, examining, understanding, and disseminating solutions to problems that are already available within the community and organizations including the healthcare system can help in bringing improvements at scale. However, to the best of the researcher’s knowledge, there is no scoping review that is conducted to map and synthesize the available evidence on the use of the PD approach for healthcare system service improvements. Hence, this scoping review aims to map and synthesize resources on the methodologies and reported outcomes and identify gaps and potentials regarding the use of the PD approach in the healthcare system.

**Methods and analysis:** Articles will be searched and retrieved in research databases such as PubMed, Embase, and Scopus. Retrieved articles will be screened independently for inclusion through a title and, or abstract review. Then, articles that passed the title and abstract review will be screened by reading the full texts. A descriptive mapping and synthesis of the literature will be employed to present data using the Preferred Reporting Items for Systematic Reviews and Meta-analysis extension for Scoping Reviews checklist and data will be presented in text, figure, and table formats.

**Ethics and dissemination:** The results of this scoping review will be published in peer-reviewed reputable international journals. Furthermore, it will also be disseminated through conference presentations, and popular press to the wider community. However, formal ethical approval is not required as primary data will not be collected.

**Strength and limitations:**

❖ This scoping review is going to be conducted in line with the updated PRISMA-SCR guideline.
❖ However, this scoping review is focused on the supply side and will only address the application of positive deviance at the health system, health facility, and health worker levels.
❖ Therefore, only the articles that have measured the performance and behavior of those entities either directly or indirectly in their therapeutic relationships with patients and the community will be included.

## Background

Achieving universal health coverage goals without the provision of high-quality healthcare services is an impossible vision (1). Quality of care is the extent to which healthcare services provided for people increase the likelihood of better health outcomes (1, 2). However, healthcare systems are facing lots of challenges in enhancing access and delivering quality healthcare services around the world including poor data collection and monitoring systems, ineffective organizational team culture and capacity limitations, ineffective leadership, failure to promote good performance (3), and lack of evidence-based health policies to assist implementation and enhance the expertise of healthcare professionals (4). Therefore, designing innovative strategies to improve healthcare delivery, including quality, has been one of the top agendas in the healthcare system in the last decades. One of the innovative strategies that has been utilized in different quality enhancement programs throughout the world is the positive deviance (PD) approach (5).

Positive deviance is an innovative approach that seeks to identify practices that result in a better outcome for the people and organizations that practice it compared with the rest of the community and other organizations (6). It is an asset-based approach, recognizing the values of prevailing expertise and experience to solve intractable issues, and is community-driven with its dynamic optimistic future having a positive effect, among others (7). The approach assumes that solutions to problems are already available within the community, and identifying and sharing those solutions can help other people identify their strengths and be prepared to resolve potential issues with the limited resources they have in their hands (6, 7). According to Jason Gordon (2022), there are always people, groups, and, or organizations who are positively deviant and who excel in their performance to rules concerning organizational issues, despite having similar constraints and resources as everyone else, and if given the opportunity, they are willing to share their experiences with other people as far as leaders are able to step back and facilitate the process (8).

Positive deviance is believed to have been introduced into the international healthcare system between the 1960s and 1970s (5, 9). A healthcare system is an organization consisting of people, institutions, and other resources, that delivers healthcare services to meet the healthcare needs of diseased individuals and the general population (10). Therefore, if the healthcare system is to ensure that the public receives the best possible health services, it has to identify and recognize people and healthcare organizations who have had exceptional performance in their healthcare practice and learn from them (11). As a result, there is a growing need to explore and use innovative approaches, as of positive deviance, to improve and bring quality healthcare services into the healthcare system arena around the world (11, 12).

According to a literature review conducted by Janine S. et al (2007), PD has been tested successfully in improving the nutritional status of children in several countries, including; Haiti, Vietnam, Pakistan, and India (13). In addition, it has also been utilized in the prevention and treatment of undernutrition, overweight, and obesity in socioeconomically vulnerable mothers and the adult population (13-15). Furthermore, several studies have used the approach in different situations, including in nursing leadership (16), in healthcare organizations (17), patient safety (18), healthcare equity in quality, and obstetric safety (19), healthcare quality and safety (20), in family planning and community-based health interventions (11, 21), and for primary care (22).

However, the available evidence on PD is mixed and there are controversies on the effectiveness of the approach, particularly in complex and demanding settings including the healthcare system. This is due in large part to methodological limitations of the extant studies including inconsistencies in the quality of strategies employed to identify PDs or positive deviant practices. For instance, it has been reported that studies that applied the PD approach for quality improvement with complex interventions lack methodological quality and details in their work (17, 23). Despite these controversies, to the best of the researchers’ knowledge, there is no scoping review that has been conducted to map and synthesize the available evidence on the methodologies, strategies utilized to identify PDs, achievements reported, and prospects of utilizing PD in healthcare systems. Therefore, this scoping review aims to map and synthesize resources and identify lessons, including successes, gaps, and potentials regarding the use of the PD approach in the healthcare system.

### Research questions

The research questions guiding this scoping review are: 1) What research designs and methods are utilized in implementing PD in the healthcare system service improvement activities?; 2) What are the strategies applied to identify positive deviants or measure positive deviant practices and identify positive deviants?; 3) What outcomes have been achieved using the PD approach?; and 4) What are the limitations in utilization of the PD approach, and what is its potential for use in the healthcare system research, and quality and safety improvement programs?

## Methods

The Preferred Reporting Items for Systematic Reviews and Meta-analysis extension for Scoping Reviews checklist (PRISMA-ScR) will be used as a guide for extraction, analysis, and presentation of the scoping review results. The PRISMA-ScR is designed to help researchers map evidence to other stakeholders including, patients or consumers, publishers, healthcare providers, policymakers, and guideline developers to have a greater understanding of relevant terminologies, core concepts, and key items to report for scoping reviews (24). The Population, Concept, and Context (PCC) framework will also be employed to assess the research question’s appropriateness. According to the PCC framework, the target population will include people of any gender and age who are considered as study participants in patient and healthcare worker therapeutic relationships, or, in any healthcare service-related studies in the health system, and the concept will be dealt with the context of utilizing a PD approach in healthcare system research. Furthermore, the context aspect will incorporate articles that took healthcare settings and health systems as part of study settings and that used the PD approach to identify high-performing individuals, teams, groups, and/or healthcare facilities, and health systems themselves, in any healthcare services, understand their strategy, and disseminate their findings to the global audience, irrespective of the location, country, and region of study.

### Literature search strategy

International databases including PubMed, Embase, Scopus, Web of Science, CINAHL, and Google Scholar will be explored to retrieve resources related to PD and its application in the healthcare system domain. Moreover, hand search and article reference tracking will also be employed to find additional literature from Google sources. Articles released through June 01, 2023, will be included, as PD is believed to be introduced into the world in the early 1970s (5, 9), and a database search will be carried out between May 15, 2023, and June 15, 2023. For duplication avoidance and citation purposes, references retrieved from each database will be stored in the EndNote desktop version x20 separately and a complete search strategy for all major database has been included as a supplementary file in the protocol and will be included as an appendix in the subsequent scoping review. In addition, the key terms and, or phrases written in table one below will be used, after being refined to fit different databases, to identify the necessary articles, and the Boolean operators of “OR” and “AND” will also be applied to combine the key terms and phrases. Furthermore, the asterisk search operator will also be used to search terms that have similar word stems or start with the same letters.

**Table 1:** Identified terms/concepts/phrases for the scoping review search strategy development.

| Theme 1 | AND | Theme 2 | AND | Theme 3 |
| --- | --- | --- | --- | --- |
| "Positive Devian*" OR |  | Method* OR |  | Health* OR |
| "Positive Deviant" OR |  | Procedure* OR |  | Healthcare* OR |
| "Positive deviance" OR |  | Technique* OR |  | "Health Care*" OR |
| "Deviance Approach" OR |  | Strateg* OR |  | "Health System*" OR |
| "Deviant Approach" OR |  | Achieve* OR |  | "Health Service*" OR |
| "Positive Outlier" OR |  | Success* OR |  | "Health Equity" OR |
| "Positive Development" |  | Accomplish* OR |  | "Quality Of Health Care* OR |
|  |  | Attain* OR |  | "Quality Improvement*" OR |
|  |  | Outcome* OR |  | "Health Behavior*" OR |
|  |  | Prospect* OR |  | "Health Promotion*" OR |
|  |  | Challenge* OR |  | "Diseases Prevention OR |
|  |  | Limit* OR |  | Treatment* OR |
|  |  | Potential* OR |  | "Palliative Care" OR |
|  |  | Opportunit* |  | "Rehabilitative Care" |

### Inclusion and exclusion criteria

Articles published only in the English language will be considered due to feasibility issues in garnering a translator and the limitation of resources. In addition, conference proceedings, oral and poster presentations, abstract articles, correspondence articles, comments to the editor, commentator, brief communication, and literature review articles other than systematic review and meta-analysis will be excluded as the scoping review requires a detailed methodological exploration and full-text results for review and data extraction. Therefore, both primary articles including observational and case studies, evaluation studies, randomized and non-randomized trials, and systematic reviews and meta-analysis articles will be included. Only peer-reviewed and published literature conducted using the PD approach in the health care system areas will be considered for reasons of credibility.

However, to be included, studies must report the methods used to measure individuals, teams, groups, or the health care facility’s strategies and interventions that are employed to bring change in the health care services including financial protection for patients, individuals, and the community at large. Therefore, community-based studies that used the PD approach to identify strategies for any health-related outcomes without the involvement of any health workers in the health system including community health workers will not be included unless it clearly shows the extent of healthcare settings and health systems involvement in the study. Because the results in a community setting may not truly reflect whether the outcome is due to the involvement of healthcare facilities, local health departments, and their staffs or not.

In addition, studies that are conducted in the health care settings at the patient level will be excluded if the results do not show that the outcomes of patient-level data are due to the healthcare services rendered by the health facilities. The reason for this is that this scoping review is designed to synthesize and map available evidence in the utilization of PD in the health care system as a supply side. However, the measures for performance of being positive deviant from the supply side can be patient outcomes including morbidity and mortality rate reductions, immunization coverage, and financial protection.

### Screening

All articles’ citations will be imported into the Covidence online tool and assessed independently for inclusion through a title and, or abstract read. Then, studies that passed the title and abstract review by the principal investigator will be screened by two independent reviewers with full-text reading. In case of duplication, the full-text published article will be retained, and others will be removed. However, articles without full access and those that did not report the outcomes of interest will be excluded.

### Data Extraction

The data extraction platform will be developed on the Excel sheet, and data examination, extraction, and sorting will be done using key themes; Author(s) name, publication year, article type, study design and method, study setting, participant type, study domain or topic, World Health Organization (WHO) geographic category/region, World Bank (WB) group and, or country, intervention, and other measures in using the PD approach for research and initiatives in all health care settings around the world. The data extraction sheet will be piloted using 5 randomly selected papers and then, the extraction form will be adjusted after having the piloted template and be utilized to customize the Covidence data extraction format 2.0. The principal investigator will independently extract all the necessary data from each article using the customized data extraction format in the Covidence online tool and then undergo a second check-up after completion of extraction from all articles. For articles that did not provide details concerning their study background, corresponding authors will be contacted through e-mail and asked for relevant information, such as study time, outcome measurement method, and other significant variables.

### Patient and public involvement

No patient or public will be involved.

### Data analysis and presentation

A descriptive mapping and synthesis of literature will be employed to present data both in text and table formats using five major themes: Methodologies used in implementing PD; criteria researchers utilized to identify positive deviants; the strategies of positive deviants that make them successful apart from others; outcomes reported from the usage of PD approach; and the prospects of PD in future health service research areas and quality improvement programs. The results of this scoping review will be reported based on the PRISMA-ScR guideline (24), and the entire process of study screening, selection, and inclusion will be shown with the support of a flow diagram. Furthermore, the PCC framework will be used to guide the presentation of the scoping review results to the wider community.

### Ethics and Dissemination

This scoping review protocol will be registered and be made freely available to the wider community by publishing it in one of the following platforms: Figshare, Open Science Framework, Research Square, ResearchGate, and, or other similar databases. However, it will not be registered in the International Prospective Register of Systematic Reviews (PROSPERO), as the database is not currently registering scoping reviews (25). The scoping review findings will be presented at conferences at The University of Queensland in Australia, the Postgraduate Institute of Medicine. In addition, it will be published in peer-reviewed, reputable international journals. Furthermore, it will also be disseminated to the wider community, through conference presentations at different international research symposiums, and popular press. However, formal ethical approval is not required as primary data will not be collected.

### Timeline for review

Start date: May 15, 2023

End date: November 30, 2023

**Stage of review:** Review ongoing

## Discussion

Achieving the universal health coverage goals without enhancing access and the provision of high-quality healthcare services is an impossible vision (1). Quality of care is the extent to which healthcare services provided for people increase the likelihood of better health outcomes (1, 2). However, delivering high-quality healthcare services is facing lots of challenges in the healthcare quality improvement programs, including poor data collection and monitoring systems, organizational team culture, and capacity limitations, leadership weakness, failure of promoting good performance (3), and lack of evidence-based health policies, to assist implementation and enhance the expertise of healthcare professionals (4). Hence, designing innovative strategies to improve healthcare service delivery, including quality, has been one of the top agendas in the healthcare system in the last decades. One of the innovative strategies that have been utilized in different healthcare services, including quality, enhancement programs throughout the world is the PD approach (5, 26). It is a strategy based on a core assumption that community members, including individuals and organizations, already have the answers to many issues and that by studying, comprehending, and disseminating these solutions to other members of the community, improvements can be made at a large scale (26). Despite this fact, evidence on the methodologies, strategies employed in selecting positive deviants, achievements in using the approach, the possible limitations and challenges of using it in practice, and its potential for future use in the healthcare system are not well documented (5, 17). In addition, the utility of the PD approach is reported to be limited in complex settings including the healthcare system in some earlier articles. For instance, Baxter R. et al (2016) stated that utilizing PD in complex and demanding healthcare settings poses challenges, citing engaging staff more broadly in quality enhancement programs as a known difficult aspect (27). However, several sources across the literature have indicated the merit of utilizing a PD approach in multiple quality improvement programs of the healthcare system including in increasing hand hygiene compliance rate (28-30), infection prevention (31, 32), reduction of postoperative complications (33), reducing medication errors (34), and improving vaccination coverage rates within healthcare systems (35). Therefore, this scoping review aims to map evidence, synthesize the available literature, and identify gaps on the use of the PD approach for health system service improvement and analyze its prospects for future healthcare quality, safety, and other service improvement programs in the healthcare system.

## Conclusion

This global scoping review protocol is designed to map and synthesize the methodologies used and strategies utilized in selecting positive deviants, achievements, and the prospects of using PD in different healthcare settings, identify the gaps, and answer the critical research questions that are developed in line with the proposed title. Moreover, the findings of the scoping review will add a body of knowledge and provide insight to policymakers and service providers regarding the opportunities of utilizing PD in healthcare system research and healthcare quality improvement programs.

## Data Availability

Not applicable

## Abbreviations

PCC: Population, Concept, and Context
PRISMA-ScR: Preferred Reporting Items for Systematic Reviews and Meta-analysis extension for Scoping Reviews
UN: United Nations
WB: World Bank
WHO: World Health Organization

## Declaration

### Consent for publication

Not applicable

### Availability of data and materials

Not applicable

## Acknowledgment

Not applicable

## Author contribution

AM and YA conceived the idea; AM planned and designed the study protocol; AM and YA planned the data extraction and statistical analysis; AM wrote the first draft. YA, EW, AZ, AE, EE, and BB reviewed the draft and provided critical insights. All authors have reviewed, participated in editions, and approved the submission of the final protocol.

## Annex 1

### PubMed Search Strategy

1. *((“Positive Devian*” OR “Positive Deviance” OR “Deviant Approach”) AND (Method* OR Procedure* OR Technique OR Strateg* OR Achieve* OR Success* OR Accomplishment* OR Attainment* OR Outcome* OR Prospect* OR Challenge* OR Limitation* OR Potential*)) AND (Health* OR Healthcare* OR “Healthcare*” OR “Health System*” OR “Health Service*” OR “Health Equity” OR “Quality Improvement*” OR “Health Behavior*” OR “Health Promotion*” OR Prevent* OR Treatment* OR “Palliative Care”)*.

Therefore, the following combinations will be employed.

❖ (“positive devian*”[All Fields] OR “Positive Deviance”[All Fields] OR ((“deviant”[All Fields] OR “deviants”[All Fields]) AND (“approach”[All Fields] OR “approach s”[All Fields] OR “approachability”[All Fields] OR “approachable”[All Fields] OR “approache”[All Fields] OR “approached”[All Fields] OR “approaches”[All Fields] OR “approaching”[All Fields] OR “approachs”[All Fields]))) AND (“method*”[All Fields] OR “procedure*”[All Fields] OR (“methods”[MeSH Terms] OR “methods”[All Fields] OR “technique”[All Fields] OR “methods”[MeSH Subheading] OR “techniques”[All Fields] OR “technique s”[All Fields]) OR “strateg*”[All Fields] OR “achieve*”[All Fields] OR “success*”[All Fields] OR “accomplishment*”[All Fields] OR “attainment*”[All Fields] OR “outcome*”[All Fields] OR “prospect*”[All Fields] OR “challenge*”[All Fields] OR “limitation*”[All Fields] OR “potential*”[All Fields]) AND (“health*”[All Fields] OR “healthcare*”[All Fields] OR “healthcare*”[All Fields] OR “health system*”[All Fields] OR “health service*”[All Fields] OR “Health Equity”[All Fields] OR “quality improvement*”[All Fields] OR “health behavior*”[All Fields] OR “health promotion*”[All Fields] OR “prevent*”[All Fields] OR “treatment*”[All Fields] OR “Palliative Care”[All Fields]). **All fields searches and without adding any filter**.

